# Better therapeutic efficiency of omnidirectional than directional subthalamic deep brain stimulation in Parkinson’s disease

**DOI:** 10.1101/2023.11.01.23297794

**Authors:** Maximilian Scherer, Luka Milosevic, Idil Cebi, Patrick Bookjans, Bastian Brunett, Robert Guggenberger, Daniel Weiss, Alireza Gharabaghi

## Abstract

**Background:** Deep brain stimulation (DBS) using segmented electrode contacts allows for directionally steered stimulation (DS), while the conventional ring mode provides omnidirectional stimulation (OS). However, with regard to achieving better effects with the same stimulation intensity or equivalent effects with lower intensity, the comparative therapeutic efficiency of these approaches remains unclear.

**Objective:** To compare the therapeutic efficiency of subthalamic DBS using segmented and ring contacts in Parkinson’s disease (PD) patients with akinetic-rigid symptoms.

**Methods:** A double-blind, randomized monopolar review was conducted with patients in the dopaminergic medication-off state at 10 weeks postoperatively on three upper ring contacts and three contacts of the upper segmented level. Impedance measurements were obtained, and the therapeutic threshold (current strength for complete biceps brachii muscle rigidity resolution) was estimated by increasing stimulation intensity in 0.2 mA increments.

**Results:** OS with ring contacts showed an improved therapeutic threshold compared to DS with segmented contacts. At 1.1 mA stimulation intensity, complete rigidity resolution was achieved in 90% of patients with the best ring contact, whereas only 40% achieved the same outcome with the best segmented contact. In addition, OS with ring contacts exhibited 50% lower impedance than DS with segmented contacts.

**Conclusions:** Incremental adjustments in current intensity during parameter titration generate valuable stimulus-response curves for assessing therapeutic efficiency. In clinical practice, the monopolar review should give priority to identifying the optimal ring level and therapeutic threshold. Segmented contacts should be carefully considered as a potential alternative when side effects limit the feasibility of other options.

## Introduction

Fluctuations in dopaminergic response in patients with idiopathic Parkinson’s disease (PD) are effectively addressed by deep brain stimulation (DBS) of the subthalamic nucleus (STN), which provides evidence-based and cost-effective treatment with superior clinical outcomes compared to the best medical treatment (Weaver et al., 2012; Schuepbach et al., 2013; Dams et al., 2016; Lhommée et al., 2018). Despite advancements in neurosurgical targeting, intraoperative techniques, awake procedures, and stimulation paradigms, a notable proportion of patients may still require revision surgery due to side effects or insufficient therapeutic benefits (Rosahl et al., 2002; Weiss et al., 2011; Millian et al., 2013; Southwell et al., 2016; Rolston et al., 2016; Naros et al., 2018; Milosevic et al., 2020).

The latest generation of DBS electrodes introduces a design modification in which the middle two ring contacts are both divided into three segments, enabling directional stimulation (DS) steering in addition to the circular or omnidirectional stimulation (OS) of the traditional ring mode (Contarino et al., 2014; Pollo et al., 2014; Steigerwald et al., 2016; Dembek et al., 2017; Schnitzler et al., 2021; Ramanathan et al., 2023; Debove et al., 2023). While simulation studies indicate that DS can reduce out-of-target stimulation and associated side effects, it may have limitations in achieving optimal therapeutic benefit when compensating for suboptimal electrode placements (Kramme et al., 2021). Recent empirical findings suggest that although DS can alleviate DBS side effects caused by misplaced electrodes, some patients may still require surgical revision (Mishra et al., 2023).

In this context, it is important to determine the potential of directional stimulation (DS) in enhancing the therapeutic efficiency of DBS in the majority of patients with well-placed electrode leads, where beneficial effects can be achieved without early side effects. However, there is still ambiguity whether DS allows for more efficient stimulation of the target structure by reducing the required stimulation intensity to achieve the desired clinical benefit compared to omnidirectional stimulation (OS) (Pollo et al., 2014; Contarino et al., 2014; Dembek et al., 2017; Schnitzler et al., 2021).

To address this open question, we performed precise stimulation amplitude titration with 0.2 mA step sizes for ring and segmented contacts, resulting in high-resolution clinical stimulation-response curves. This facilitated rigorous comparisons of DS and OS effects at various stimulation intensities, primarily establishing the threshold for resolving muscular rigidity in PD patients. Specifically, ten weeks after DBS lead implantation, a randomized, double-blind monopolar review was conducted on the three upper contact levels and the three parts of the upper segmented level. Impedance measurements were also obtained for each contact to assess potential clinical benefits in relation to power consumption and battery life.

Our hypothesis was that spatially focused DS of the STN would lead to more efficient resolution of upper limb rigidity compared to OS, i.e., better effects would be achieved with the same stimulation intensity or equivalent effects with lower intensity.

## Methods

### Patients

The akinetic-rigid PD patients of this study had participated in the SANTOP study (“Subthalamic Steering for Therapy Optimization in Parkinson’s Disease”; ClinicalTrials.gov: NCT03548506) that evaluated the long-term effects of omnidirectional vs. directional deep brain stimulation in a randomized, cross-over protocol six months after surgery; the respective results are reported elsewhere (Gharabaghi et al., 2023). Here, we report the findings of a monopolar review conducted ten weeks after surgery on two consecutive days, i.e., evaluating the three upper ring contacts (day 1) and the three segments of the upper segmented level (day 2). The complete data for analysis was available in seventeen of twenty-one participating PD patients, in whom the rigidity of the right biceps brachii muscle was assessed during DBS of the left subthalamic nucleus in the dopaminergic medication-off state. Furthermore, impedances were measured for all contacts in every patient.

Each patient underwent bilateral implantation of STN-DBS leads (6170, Abbott laboratories, Lake Bluff, Illinois, U.S.) on average 69 days prior to the evaluation. The implantation target was preoperatively identified via magnetic resonance imaging (MRI) and computer tomography (CT) images. Intraoperatively, successful implantation was validated by local field potential beta-peaks (Milosevic et al., 2020) or microelectrode recordings (Hutchison et al., 1998; Knieling et al., 2016). Postoperative CT images were co-registered with preoperative MRI to confirm accurate electrode implantation. Patients were examined several weeks after the DBS lead implantation to avoid micro-lesion effects (Granziera et al., 2008), and after overnight withdrawal from dopaminergic medication. Written informed consent was provided by all patients and the study was approved by the ethics committee of the Medical Faculty of Tübingen. Detailed information on patient demographics is provided in Table 1.

**Table 1:**
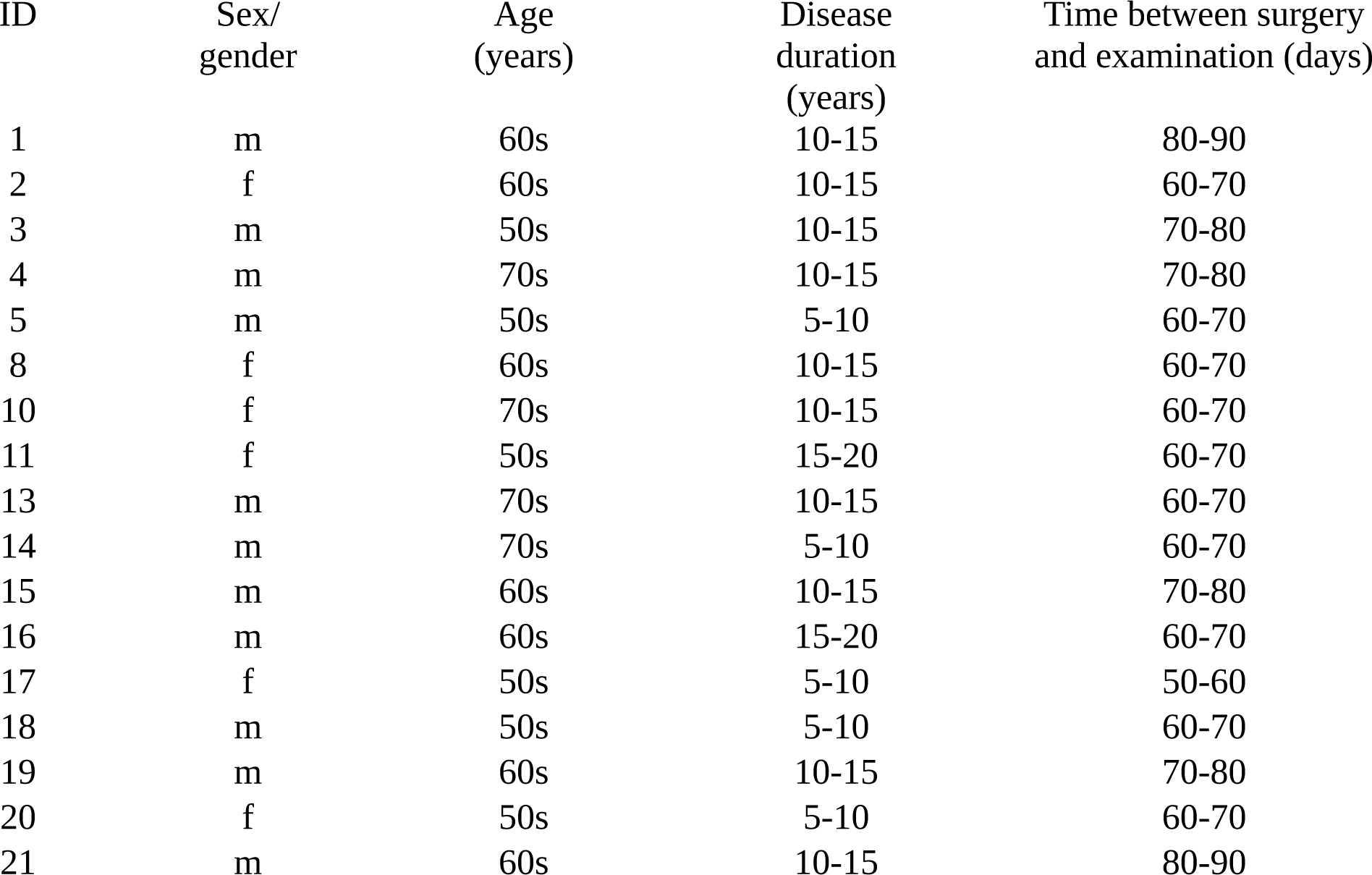
Patient information.

### Stimulation configuration

The three upper ring contacts were evaluated in all seventeen patients. In addition, the three segments of the upper segmented level were investigated in sixteen patients, and of the lower segmented level in one patient. Stimulation was always applied at 130 Hz and 60 µs while increasing the stimulation intensity by 0.2 mA stepwise (see experimental protocol). Stimulation was applied to the left STN while assessing the rigidity of the right arm.

### Data acquisition and experimental protocol

The patients were instructed to relax their arms and to remain awake. Each run lasted 90 seconds and comprised two phases; a non-movement phase (30 s) and a continuous passive movement phase (60 s). During the continuous passive movement phase, an examiner moved the subject’s arm at a frequency of 0.5 Hz (acoustically communicated to the examiner via headphones). At the end of each run, the examiner provided an estimate of the patient’s rigidity according to the Unified Parkinson’s Disease Ratings Scale (MDS-UPDRS) assessment. For later analyses, these scores were binarized to represent either a lack (UPDRS score of zero) or the presence of rigidity. In the first run, patients were evaluated at OFF stimulation. In the consecutive runs, stimulation was increased from 0.5 mA to 2.5 mA in 0.2 mA increments on one stimulation contact before switching to another contact. The contacts were evaluated in randomized order. Before switching contacts, the paradigm was paused for two minutes to avoid stimulation-related carry-over effects (Levin et al., 2009). Above 2.5 mA, the evaluation was continued in 0.2 mA increments, albeit the evaluation was modified to identify side effects, i.e., without concurrent arm movements. Due to the fact that the programming was performed by a third person, the patient and the examiner were unaware as to which stimulation contact was evaluated at which intensity.

### Determining the therapeutic threshold

The therapeutic threshold was identified for each ring/segmented contact in each patient. It was defined as the minimal stimulation intensity from which on rigidity remained completely resolved. Specifically, only consistent resolving of rigidity was considered to be the therapeutic threshold. For example, if rigidity was completely resolved at 0.9 mA and 1.3 mA and all higher intensities, but was still present at intensities of 0.7 mA and 1.1 mA; the therapeutic threshold was determined at 1.3 mA.

### Determining the side-effect threshold

The side-effect threshold was also identified for each ring/segmented contact in each patient. It was defined as the lowest stimulation intensity which elicited side-effects such as muscle cramps or double vision.

### Determining therapeutic window size

The therapeutic window was determined as the difference between the lower (i.e., therapeutic threshold) and upper (i.e., side-effect threshold) border of clinical efficacy.

### Determining contact impedance

The impedance values were measured intraoperatively, i.e., via the integrated functionality of the programming device, by attaching an external pulse generator at the end of the surgery. Low intensity bipolar stimulation was utilized to measure the impedance between individual contacts. The impedance of a specific contact was calculated as the average impedance between it and the other contacts.

### Statistical evaluations

For statistical evaluation, the contacts were classified on the basis of different categories: (i) according to level, i.e., ring 4 (most upper), ring 3 (second most upper), ring 2 (third most upper), frontal segment, medial segment, lateral segment; (ii) best, second best, worst ring/segment with regard to the therapeutic threshold; (iii) best, second best, worst ring/segment with regard to the side-effect threshold; (iv) best, second best, worst ring/segment with regard to the therapeutic window size (as the range between therapeutic threshold and side-effect threshold). The stimulation effect was quantified using a linear mixed model. The measured clinical efficacy was modeled as the dependent variable. The other variables were modeled as independent variables: Stimulation intensity was included as a categorical fixed factor to estimate the effect of stimulation OFF versus a specific stimulation intensity; the patient ID was included as a categorical random factor to compensate for repeated measurements over individual patients.

Patient-wise paired t-tests were applied to investigate the differences between the therapeutic thresholds, side effect thresholds and therapeutic window sizes of ring and segmented contacts. Multiple comparison correction (MCC) was applied using the Bonferroni method, an alpha value of 0.05 and a hypothesis count matching the count of all statistical evaluations presented in this work. The results are presented with a marker indicating either significance or non-significance after MCC.

### Code and data accessibility

Data and evaluation code will be shared by the first author upon request. The toolbox FiNN (Scherer et al., 2022) that was used to analyze the data is available on GitHub.

## Results

The ratio of rigid trials decreased consistently with increasing stimulation intensity. This result was independent of grouping of the contacts on the basis of different categories, i.e., according to the level (2, 3, or 4) of the stimulated electrode (Figure 1 A), therapeutic window size (Figure 1 B, C), therapeutic threshold (Figure 2 A, B), and side-effect threshold (Figure 2 C, D).

**Figure 1:**
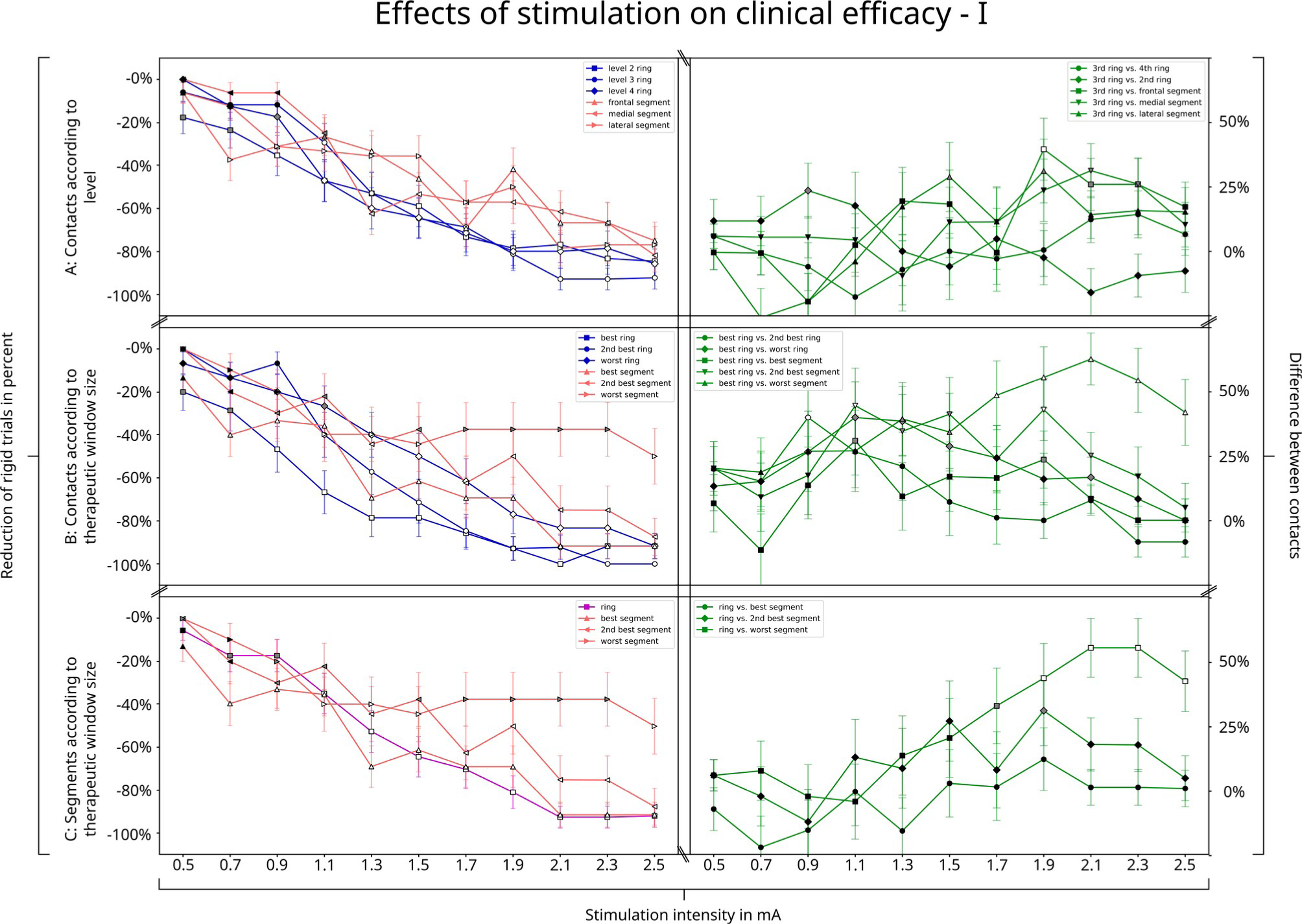
Effects of stimulation on clinical efficacy – I. Subfigure A shows the findings for contacts grouped by level (spatial position), subfigure B for contacts grouped by therapeutic window size and subfigure C for segmented contacts ranked by therapeutic threshold in comparison to the corresponding ring mode of the same segments. White, gray and black colors of the symbols indicate significant (after MCC), significant (before MCC) and non-significant (i) reduction of rigid trials across all patients (plots on the left side), and (ii) differences between contacts (green plots on the right side), respectively. Specifically, 40% reduction of rigid trials at 0.7 mA with the best segmented contact indicated that 40% of patients had complete resolving of rigidity at this specific contact and intensity; a result that was significant after MCC in comparison to baseline without stimulation. However, when comparing this effect to the effects of the other contacts at the same stimulation intensity (see plots on the right side), no significant difference was found.

**Figure 2:**
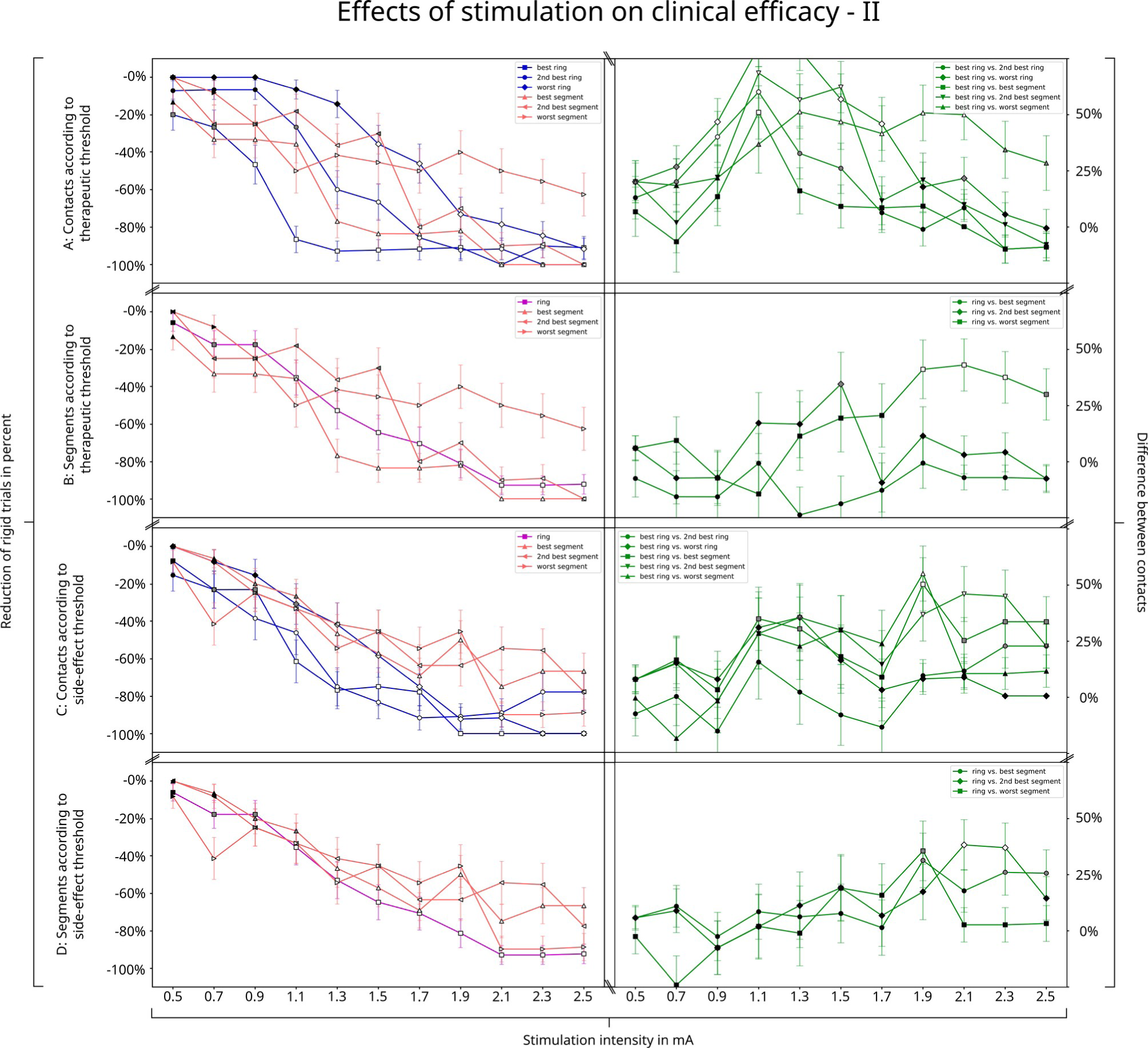
Effects of stimulation on clinical efficacy – II. Same as figure 1 but for contacts classified by therapeutic threshold (subfigures A and B), and side-effect threshold (subfigures C and D)

In the following, we explain by way of example how the individual plots can be set in relation to each other: When stimulating at an intensity of 0.7 mA, the ratio of rigid trials was significantly reduced for one contact in comparison to baseline (i.e., without stimulation); at this intensity, 40% of patients had complete resolving of rigidity when stimulated via a laterally steering segmented contact (Figure 1 A). At the intensity of 0.7 mA, also 40% of patients had the best therapeutic window (Figure 1 B, C), the best therapeutic threshold (Figure 2 A, B), and worst side-effect threshold (Figure 2 C, D). This may suggest that, at 0.7 mA, laterally steering segments may present large therapeutic windows via an improved therapeutic threshold. The stimulation effect of the laterally steering contact plateaued from 2.1 mA onwards, and achieved complete resolving of rigidity in 80% of the patients. A comparison of these effects with the other contacts (both segmented and ring) at the same stimulation intensity did not reveal any significant differences (see respective plots on the right side of Figures 1 and 2).

When stimulated at 1.3 mA, all contacts (both ring and segmented) showed a significant reduction of rigidity in comparison to baseline (i.e., stimulation off); the second-highest ring contact led to the largest reduction of rigidity in comparison to baseline, plateaued from 2.1 mA onwards, and achieved complete resolving of rigidity in 90% of the patients (Figure 1 A). However, a comparison of these effects with the other contacts (both segmented and ring) at the same stimulation intensity did not reveal any significant differences (see respective plots on the right side of Figures 1 and 2).

Notably, the ring contact with the lowest therapeutic threshold achieved complete resolution of rigidity at 1.1 mA (and plateaued onwards) in 90% of the patients (Figure 2 A, left). This was significantly better than the segmented contact with the lowest therapeutic threshold, where 40% of the patients had complete resolving of rigidity (Figure 2 A, right). However, a further increase in stimulation intensity did not cause any significant difference between the best ring and the other contacts (apart from the worst segmented contact).

This suggests that there is an optimal stimulation intensity for complete resolution of rigor for the best ring contact. The best ring contact with the therapeutic threshold at 1.1 mA (Figure 2 A), was usually (but not always) the contact with the largest therapeutic window (Figure 1 B) and the highest side-effect threshold (Figure 2 C), but could not be attributed to a single electrode level (Figure 1 A).

With regard to the therapeutic threshold, a comparison between the segmented contacts and the corresponding ring mode (Figure 2 B) did not reveal any significant differences, thus indicating that the selection of the optimal ring along the implantation trajectory had a larger influence on rigidity than directional steering (Figure 2 A). This observation is supported by the comparison between the therapeutic windows of ring and segmented electrodes:

Specifically, with regard to the therapeutic window the best ring contact was significantly larger (Figure 3A) than the second best (p < 0.05) and worst ring contact (p < 0.01). A comparison between the segments and their corresponding ring mode with regard to the therapeutic window (Figure 3 B) showed a significant difference for the clinically worst segmented contact only (p < 0.05).

**Figure 3:**
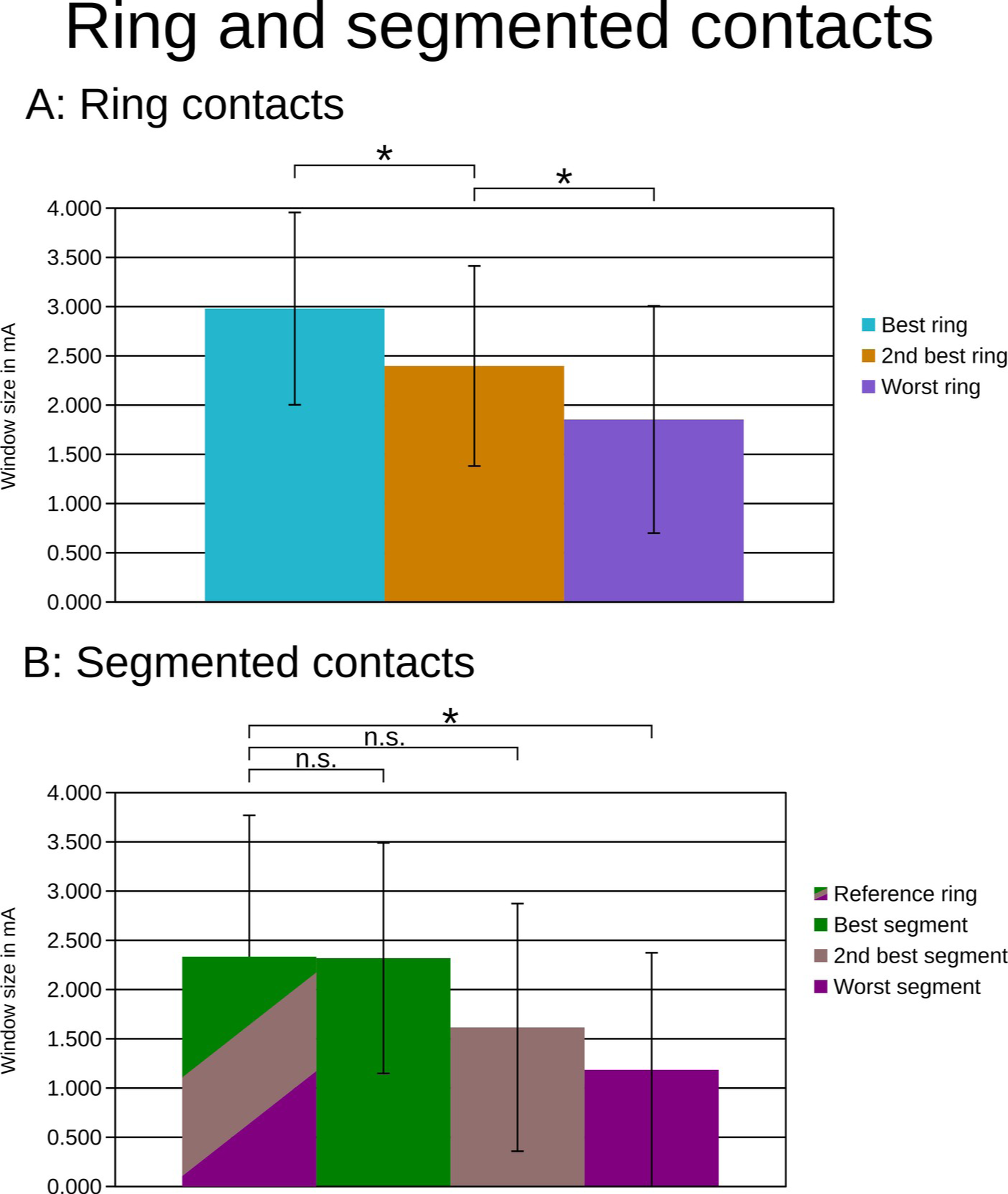
Therapeutic window of ring and segmented contacts. A: The best ring contact provided a larger therapeutic window than the other two ring contacts. B: The therapeutic window of the segmented contacts and the corresponding ring contact, i.e., the sum of the three segments, did not differ significantly, except for the worst segment.

The therapeutic window sizes were 2.3 mA (1.4 mA – 3.7 mA), 2.3 mA (1.3 mA – 3.6 mA), 1.6 mA (1.8 mA – 3.4 mA) and 1.2 mA (2.0 mA – 3.2 mA), for the ring mode; best, second best and worst segmented contacts, respectively. Stimulating via segmented contacts did not improve the therapeutic window in comparison to the respective ring mode (Figure 3 B), but doubled the impedance (p < 0.001; Figure 4). This indicates that directional stimulation did not improve the treatment-relevant stimulation thresholds and would, when applied, chronically necessitate higher power consumption than ring mode due to increased impedances.

**Figure 4:**
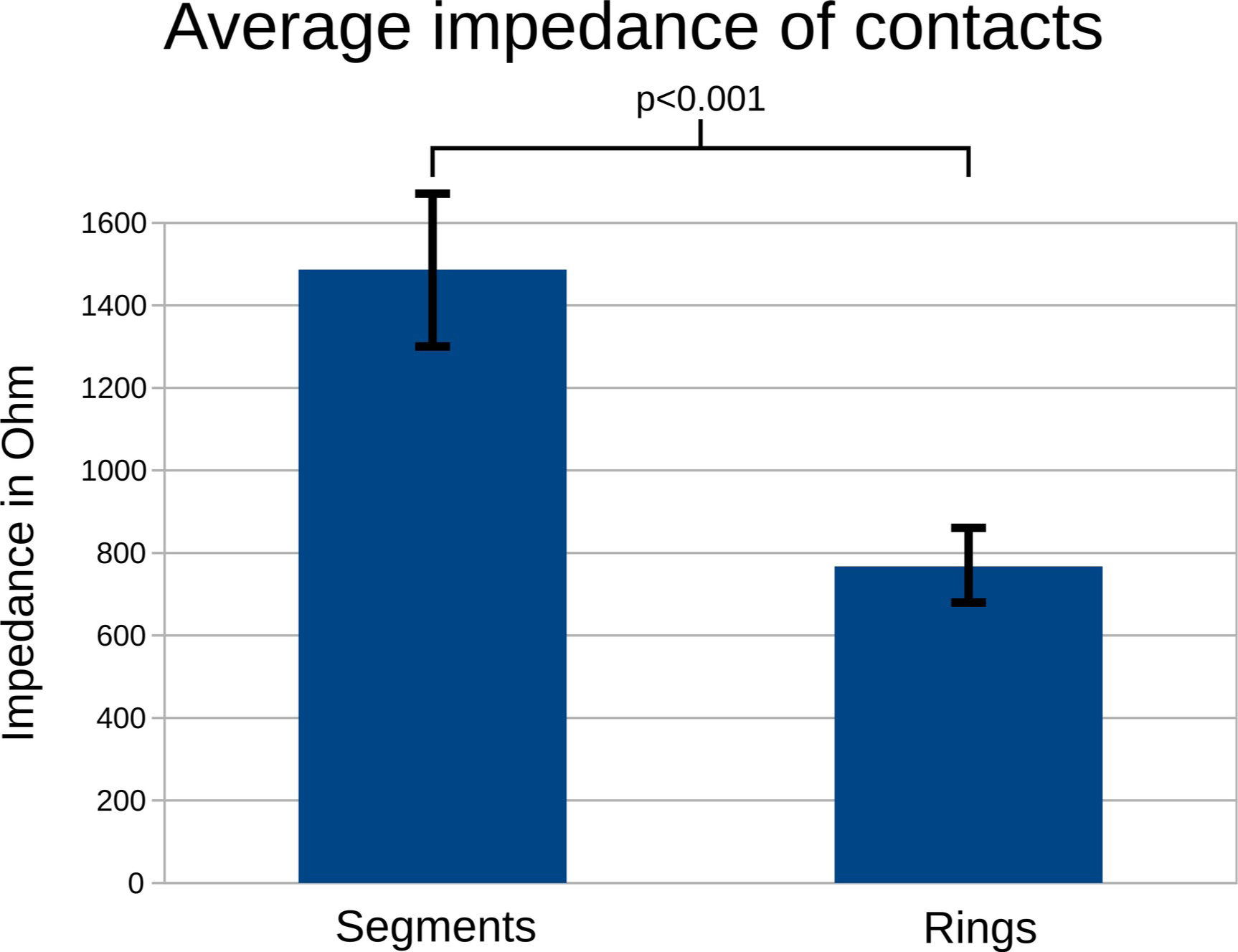
Average impedance of contacts. The impedance of segmented contacts was twice as high as for ring contacts, thus necessitating a higher energy delivery when the same stimulation parameters were applied.

## Discussion

The present study yielded new insights with regard to the efficiency of different deep brain stimulation (DBS) approaches in treating muscular rigidity in Parkinson’s disease (PD) patients. Contrary to previous research, it was determined that omnidirectional stimulation (OS) with ring contacts outperformed directional stimulation (DS) with segmented contacts, showing greater effects at the same intensity while consuming less energy.

### Therapeutic efficiency

To determine therapeutic efficiency, a high-resolution threshold detection method involving small-stepped titration (0.2 mA) was employed during the DBS monopolar review ten weeks after implantation. This method was applied to both segmented and ring contacts, which had been previously investigated using larger step sizes of 0.5 mA (Steigerwald et al., 2016) or 1.0 mA (Dembeck et al., 2017) in other studies. This approach allowed for the identification of the optimal therapeutic threshold, representing the minimum electrical current required for maximum clinical effect.

Remarkably, at the lowest therapeutic threshold complete resolving of rigidity was achieved in 90% of patients using OS with the best ring contact, at a stimulation intensity of 1.1 mA. In contrast, DS with the segmented contact with the lowest therapeutic threshold achieved complete resolving of rigidity in only 40% of patients at the same stimulation intensity.

### Vertical vs. horizontal plane

Additionally, the present study revealed that the therapeutic window (which also accounts for the side-effect threshold) of the best ring contact was significantly larger than that of the other ring contacts. Contrary to previous reports (Contarino et al., 2014; Pollo et al., 2014; Dembek et al., 2017; Schnitzler et al., 2021) however, we ascertained that stimulation with the segmented contact with the largest therapeutic window did not enhance the therapeutic window, unlike switching between different ring contacts.

While the choice of the ring contact had the largest effect on the therapeutic threshold, switching to segmented stimulation was associated with an unchanged or worsened therapeutic window size and side-effect threshold. Interestingly, the optimal ring contact could not be attributed to a specific electrode level, indicating that determining the best stimulation level along the vertical implantation trajectory was more crucial than identifying the best direction in the horizontal plane.

### Threshold detection

The results of this study diverge from previous reports, suggesting that DS exhibits a larger therapeutic window than OS with ring contacts (Contarino et al., 2014; Pollo et al., 2014; Dembek et al., 2017; Schnitzler et al., 2021). These earlier studies associated DS with either an increased side-effect threshold (Contarino et al., 2014; Dembek et al., 2017) or a decreased therapeutic threshold (Pollo et al., 2014; Schnitzler et al., 2021). However, some studies observed no improvements in therapeutic threshold (Contarino et al., 2014; Dembek et al., 2017) or therapeutic window (Steigerwald et al., 2016; Debove et al., 2023) when comparing DS to OS. These contradictory findings in prior research may be partially attributed to differences in the applied stimulation titration step sizes for DS and OS.

Due to the higher surface current density of smaller segmented contacts in comparison to that of larger ring contacts, DS is more sensitive to stimulation amplitude adjustments. It requires smaller amplitude increments (0.1-0.3 mA) than the traditional 0.5 mA used for OS to achieve a similar threshold detection sensitivity. However, if such small-stepped assessment is employed for DS, it should also be applied to OS as there is no drawback from selecting small step sizes also for OS.

One of the studies reporting a difference in therapeutic threshold between DS and OS in PD patients did not specify the stimulation titration protocol (Schnitzler et al., 2021). The other study utilized titration step sizes of 0.1 mA for both DS and OS during intraoperative assessments (Pollo et al., 2014). In view of the fact that our study employed titration step sizes of 0.2 mA for both DS and OS, this approach appears to be sufficiently sensitive for identifying threshold differences during a postoperative monopolar review.

### Power consumption

Furthermore, the smaller stimulating surface area and higher impedance of segmented contacts result in a higher total electrical energy being delivered (TEED) during stimulation compared to ring contacts (Merola et al., 2021). This increased energy consumption leads to more frequent battery replacements for non-rechargeable impulse generators (IPG), thereby posing an increased risk of morbidity due to the potential for surgical site infections (Fakhar et al., 2013; Ondo et al., 2007). The likelihood of infections rises as the number of IPG replacements increases (Pepper et al., 2013).

To address this issue, we measured the impedance of both ring and segmented contacts and ascertained that the impedance of segmented contacts was, on average, twice as high as that of ring contacts, which was consistent with earlier studies (Eleopra et al., 2019). This higher impedance leads to increased power consumption by the IPG when applied chronically, since more electrical energy is delivered (Koss et al., 2005).

Consequently, to maintain battery longevity, it is necessary to achieve the same stimulation effects with a TEED equivalent to that of OS. This would require approximately 30% less stimulation intensity for DS than for OS (Rebelo et al., 2018). However, even when applying the same stimulation intensity, such as 1.1 mA, DS proved to be either equally or less effective than OS in resolving muscular rigidity.

### Limitations and considerations

Despite our efforts to ensure a systematic comparison between DS and OS, this study has certain limitations. Firstly, although our sample size is larger than that of other single-center studies, it remains considerably smaller than a recent multicenter study (Schnitzler et al., 2021). However, one major strength of our study is the rigorous procedure employed for parameter titration in each patient, with small increments in current intensity for both stimulation conditions, resulting in consistent stimulus-response curves.

Furthermore, to streamline the assessments and minimize examination time for the patients, we made several concessions that may have reduced the sensitivity of the evaluation. Stimulation intensities were increased in step sizes of 0.2 mA instead of 0.1 mA (Pollo et al., 2014). The assessment of rigidity was limited to a maximum intensity of 2.5 mA, as we anticipated reaching the therapeutic threshold at this stimulation intensity. In addition, evaluations of segmented electrodes were restricted to one level.

Moreover, the examinations conducted in this study focused solely on the resolving of rigidity and the acute side effects induced by stimulation. Potential effects, both positive and negative, of DBS on other motor or non-motor symptoms such as cognition were not investigated. Finally, the findings presented in acute assessments need to be complemented by long-term clinical evaluations to establish reliable comparisons between OS and DS (Gharabaghi et al., 2023).

## Conclusion

By employing incremental increases in stimulation amplitude in steps of 0.2 mA during the monopolar review, it becomes feasible to discern the disparities in therapeutic threshold between OS and DS. Our findings demonstrate that the therapeutic threshold is more dependent on ring contact level selection than steering through segmented contacts. Notably, at a stimulation intensity of 1.1 mA, complete resolution of rigidity was achieved in 90% of patients with the best ring contact while concurrently necessitating lower power consumption than DS. Based on the presented insights, we suggest that the monopolar review should prioritize the identification of the optimal ring level and therapeutic threshold. Segmented contacts should only be cautiously considered as an exceptional recourse when side effects preclude alternative options.

## Credit authorship contribution statement

Maximilian Scherer: Methodology, Formal analysis, Writing – original draft.

Luka Milosevic: Formal analysis, Writing – review & editing.

Idil Cebi: Investigation, Writing – review & editing.

Patrick Bookjans: Investigation, Writing – review & editing.

Bastian Brunnett: Investigation, Writing – review & editing.

Robert Guggenberger: Formal analysis, Writing – review & editing.

Daniel Weiss: Conceptualization, Writing – review & editing.

Alireza Gharabaghi: Conceptualization, Funding acquisition, Project administration, Writing – original draft.

## Funding

This investigator-initiated trial was supported by Abbott / St. Jude Medical (SANTOP). The funding had no impact on the study design, on the collection, analysis and interpretation of data, on the writing of the report or on the decision to submit the article for publication.

## Acknowledgements

A.G. was supported by the German Federal Ministry of Education and Research (BMBF). M.S. was supported by the Alexander von Humboldt Foundation. D.W. was supported by the German Research Council (WE5375/1-3) and the Michael J Fox Foundation. We acknowledge support by the Open Access Publishing Fund of the University of Tübingen.

## Declarations of competing interests

A.G. was supported by research grants from Medtronic, Abbott, Boston Scientific, all of which were unrelated to this work. D. W. was supported by travel grants, speaker honoraria and research grants from Abbott, Abbvie, Bial, Boston Scientific, Medtronic, Kyowa Kirin, Stadapharm, all of which were unrelated to this work.

## Notes

### Clinical Trial

NCT03548506

### Author Declarations

Ethics committee of the Medical Faculty of Tuebingen, Germany gave ethical approval for this work

## Literature

Contarino, M.F., Bour, L.J., Verhagen, R., Lourens, M.A.J., de Bie, R.M.A., van den Munckhof, P., Schuurman, P.R., 2014. Directional steering. Neurology 83, 1163. 10.1212/WNL.0000000000000823

Dams, J., Balzer-Geldsetzer, M., Siebert, U., Deuschl, G., Schuepbach, W.M.M., Krack, P., Timmermann, L., Schnitzler, A., Reese, J.-P., Dodel, R., EARLYSTIM-investigators, 2016. Cost-effectiveness of neurostimulation in Parkinson’s disease with early motor complications. Mov Disord 31, 1183–1191. 10.1002/mds.26740

Debove, I., Petermann, K., Nowacki, A., Nguyen, T.-A.K., Tinkhauser, G., Michelis, J.P., Muellner, J., Amstutz, D., Bargiotas, P., Fichtner, J., Schlaeppi, J.A., Krack, P., Schuepbach, M., Pollo, C., Lachenmayer, M.L., 2023. Deep Brain Stimulation: When to Test Directional? Mov Disord Clin Pract 10, 434–439. 10.1002/mdc3.13667

Dembek, T.A., Reker, P., Visser-Vandewalle, V., Wirths, J., Treuer, H., Klehr, M., Roediger, J., Dafsari, H.S., Barbe, M.T., Timmermann, L., 2017. Directional DBS increases side-effect thresholds—A prospective, double-blind trial. Movement Disorders 32, 1380–1388. 10.1002/mds.27093

Eleopra, R., Rinaldo, S., Devigili, G., Lettieri, C., Mondani, M., D’Auria, S., Piacentino, M., Pilleri, M., 2019. Brain impedance variation of directional leads implanted in subthalamic nuclei of Parkinsonian patients. Clinical Neurophysiology 130, 1562– 1569. 10.1016/j.clinph.2019.06.001

Fakhar, K., Hastings, E., Butson, C.R., Foote, K.D., Zeilman, P., Okun, M.S., 2013. Management of Deep Brain Stimulator Battery Failure: Battery Estimators, Charge Density, and Importance of Clinical Symptoms. PLOS ONE 8, e58665. 10.1371/journal.pone.0058665

Gharabaghi A, Cebi I, Scherer M, Bookjans P, Brunnett B, Milosevic L,Weiss D, 2023. Long-term effects of directional deep brain stimulation in Parkinson’s disease: a randomized clinical trial on motor and non-motor symptoms. medRxiv. doi: 10.1101/2023.10.30.23297793

Granziera, C., Pollo, C., Russmann, H., Staedler, C., Ghika, J., Villemure, J.-G., Burkhard, P.R., Vingerhoets, F.J.G., 2008. Sub-acute delayed failure of subthalamic DBS in Parkinson’s disease: The role of micro-lesion effect. Parkinsonism & Related Disorders 14, 109–113. 10.1016/j.parkreldis.2007.06.013

Hutchison, W.D., Allan, R.J., Opitz, H., Levy, R., Dostrovsky, J.O., Lang, A.E., Lozano, A.M., 1998. Neurophysiological identification of the subthalamic nucleus in surgery for Parkinson’s disease. Annals of Neurology 44, 622–628. 10.1002/ana.410440407

Knieling, S., Sridharan, K.S., Belardinelli, P., Naros, G., Weiss, D., Mormann, F., Gharabaghi, A., 2016. An Unsupervised Online Spike-Sorting Framework. Int J Neural Syst 26, 1550042. 10.1142/S0129065715500422

Koss, A.M., Alterman, R.L., Tagliati, M., Shils, J.L., 2005. Calculating total electrical energy delivered by deep brain stimulation systems. Annals of Neurology 58, 168–168. 10.1002/ana.20525

Kramme, J., Dembek, T.A., Treuer, H., Dafsari, H.S., Barbe, M.T., Wirths, J., Visser-Vandewalle, V., 2020. Potentials and Limitations of Directional Deep Brain Stimulation: A Simulation Approach. Stereotactic and Functional Neurosurgery 99, 65–74. 10.1159/000509781

Levin, J., Krafczyk, S., Valkovič, P., Eggert, T., Claassen, J., Bötzel, K., 2009. Objective measurement of muscle rigidity in parkinsonian patients treated with subthalamic stimulation. Movement Disorders 24, 57–63. 10.1002/mds.22291

Lhommée, E., Wojtecki, L., Czernecki, V., Witt, K., Maier, F., Tonder, L., Timmermann, L., Hälbig, T.D., Pineau, F., Durif, F., Witjas, T., Pinsker, M., Mehdorn, M., Sixel-Döring, F., Kupsch, A., Krüger, R., Elben, S., Chabardès, S., Thobois, S., Brefel-Courbon, C., Ory-Magne, F., Regis, J.-M., Maltête, D., Sauvaget, A., Rau, J., Schnitzler, A., Schüpbach, M., Schade-Brittinger, C., Deuschl, G., Houeto, J.-L., Krack, P., EARLYSTIM study group, 2018. Behavioural outcomes of subthalamic stimulation and medical therapy versus medical therapy alone for Parkinson’s disease with early motor complications (EARLYSTIM trial): secondary analysis of an open-label randomised trial. Lancet Neurol 17, 223–231. 10.1016/S1474-4422(18)30035-8

Merola, A., Singh, J., Reeves, K., Changizi, B., Goetz, S., Rossi, L., Pallavaram, S., Carcieri, S., Harel, N., Shaikhouni, A., Sammartino, F., Krishna, V., Verhagen, L., Dalm, B., 2021. New Frontiers for Deep Brain Stimulation: Directionality, Sensing Technologies, Remote Programming, Robotic Stereotactic Assistance, Asleep Procedures, and Connectomics. Frontiers in Neurology 12.

Milian, M., Luerding, R., Ploppa, A., Decker, K., Psaras, T., Tatagiba, M., Gharabaghi, A., Feigl, G.C., 2013. “Imagine your neighbor mows the lawn”: a pilot study of psychological sequelae due to awake craniotomy: clinical article. J Neurosurg 118, 1288–1295. 10.3171/2013.2.JNS121254

Milosevic, L., Scherer, M., Cebi, I., Guggenberger, R., Machetanz, K., Naros, G., Weiss, D., Gharabaghi, A., 2020. Online Mapping With the Deep Brain Stimulation Lead: A Novel Targeting Tool in Parkinson’s Disease. Movement Disorders 35, 1574–1586. 10.1002/mds.28093

Mishra, A., Unadkat, P., McBriar, J.D., Schulder, M., Ramdhani, R.A., 2023. An Institutional Experience of Directional Deep Brain Stimulation and a Review of the Literature. Neuromodulation S1094–7159(22)01408–8. 10.1016/j.neurom.2022.12.008

Naros, G., Grimm, F., Weiss, D., Gharabaghi, A., 2018. Directional communication during movement execution interferes with tremor in Parkinson’s disease. Mov Disord 33, 251–261. 10.1002/mds.27221

Ondo, W.G., Meilak, C., Vuong, K.D., 2007. Predictors of battery life for the Activa® Soletra 7426 Neurostimulator. Parkinsonism & Related Disorders 13, 240–242. 10.1016/j.parkreldis.2006.11.002

Pepper, J., Zrinzo, L., Mirza, B., Foltynie, T., Limousin, P., Hariz, M., 2013. The Risk of Hardware Infection in Deep Brain Stimulation Surgery Is Greater at Impulse Generator Replacement than at the Primary Procedure. SFN 91, 56–65. 10.1159/000343202

Pollo, C., Kaelin-Lang, A., Oertel, M.F., Stieglitz, L., Taub, E., Fuhr, P., Lozano, A.M., Raabe, A., Schüpbach, M., 2014. Directional deep brain stimulation: an intraoperative double-blind pilot study. Brain 137, 2015–2026. 10.1093/brain/awu102

Ramanathan, P.V., Salas-Vega, S., Shenai, M.B., 2023. Directional Deep Brain Stimulation-A Step in the Right Direction? A Systematic Review of the Clinical and Therapeutic Efficacy of Directional Deep Brain Stimulation in Parkinson Disease. World Neurosurg 170, 54–63.e1. 10.1016/j.wneu.2022.11.085

Rebelo, P., Green, A.L., Aziz, T.Z., Kent, A., Schafer, D., Venkatesan, L., Cheeran, B., 2018. Thalamic Directional Deep Brain Stimulation for tremor: Spend less, get more. Brain Stimulation 11, 600–606. 10.1016/j.brs.2017.12.015

Rolston, J.D., Englot, D.J., Starr, P.A., Larson, P.S., 2016. An unexpectedly high rate of revisions and removals in deep brain stimulation surgery: analysis of multiple databases. Parkinsonism & related disorders 33, 72–77.

Rosahl, S.K., Gharabaghi, A., Liebig, T., Feste, C.D., Tatagiba, M., Samii, M., 2002. Skin markers for surgical planning for intradural lesions of the thoracic spine. Technical note. Surg Neurol 58, 346–348. 10.1016/s0090-3019(02)00863-7

Scherer, M., Milosevic, L., Guggenberger, R., Maus, V., Naros, G., Grimm, F., Bucurenciu, I., Steinhoff, B.J., Weber, Y.G., Lerche, H., Weiss, D., Rona, S., Gharabaghi, A., 2020. Desynchronization of temporal lobe theta-band activity during effective anterior thalamus deep brain stimulation in epilepsy. NeuroImage 218, 116967. 10.1016/j.neuroimage.2020.116967

Scherer, M., Wang, T., Guggenberger, R., Milosevic, L., Gharabaghi, A., 2022. FiNN: A toolbox for neurophysiological network analysis. Network Neuroscience 1–34. 10.1162/netn_a_00265

Schnitzler, A., Mir, P., Brodsky, M.A., Verhagen, Leonard, Groppa, S., Alvarez, R., Evans, A., Blazquez, M., Nagel, S., Pilitsis, J.G., Pötter-Nerger, M., Tse, W., Almeida, L., Tomycz, N., Jimenez-Shahed, J., Libionka, W., Carrillo, F., Hartmann, C.J., Groiss, S.J., Glaser, M., Defresne, F., Karst, E., Cheeran, B., Vesper, J., Schnitzler, A., Vesper, J., Mir, P., Verhagen, Leonardo, Tomcyz, N., Hartmann, C.J., Groppa, S., Alvarez, R., Pilitsis, J., Pötter-Nerger, M., Groiss, S.J., Brodsky, M.A., 2022. Directional Deep Brain Stimulation for Parkinson’s Disease: Results of an International Crossover Study With Randomized, Double-Blind Primary Endpoint. Neuromodulation 25, 817–828. 10.1111/ner.13407

Schuepbach, W. m. m., Rau, J., Knudsen, K., Volkmann, J., Krack, P., Timmermann, L., Hälbig, T. d., Hesekamp, H., Navarro, S. m., Meier, N., Falk, D., Mehdorn, M., Paschen, S., Maarouf, M., Barbe, M. t., Fink, G. r., Kupsch, A., Gruber, D., Schneider, G.-H., Seigneuret, E., Kistner, A., Chaynes, P., Ory-Magne, F., Brefel Courbon, C., Vesper, J., Schnitzler, A., Wojtecki, L., Houeto, J.-L., Bataille, B., Maltête, D., Damier, P., Raoul, S., Sixel-Doering, F., Hellwig, D., Gharabaghi, A., Krüger, R., Pinsker, M. o., Amtage, F., Régis, J.-M., Witjas, T., Thobois, S., Mertens, P., Kloss, M., Hartmann, A., Oertel, W. h., Post, B., Speelman, H., Agid, Y., Schade-Brittinger, C., Deuschl, G., 2013. Neurostimulation for Parkinson’s Disease with Early Motor Complications. N Engl J Med 368, 610–622. 10.1056/NEJMoa1205158

Southwell, D.G., Narvid, J.A., Martin, A.J., Qasim, S.E., Starr, P.A., Larson, P.S., 2016. Comparison of Deep Brain Stimulation Lead Targeting Accuracy and Procedure Duration between 1.5- and 3-Tesla Interventional Magnetic Resonance Imaging Systems: An Initial 12-Month Experience. Stereotactic and Functional Neurosurgery 94, 102–107. 10.1159/000443407

Steigerwald, F., Müller, L., Johannes, S., Matthies, C., Volkmann, J., 2016. Directional deep brain stimulation of the subthalamic nucleus: A pilot study using a novel neurostimulation device. Movement Disorders 31, 1240–1243. 10.1002/mds.26669

Weaver, F.M., Follett, K.A., Stern, M., Luo, P., Harris, C.L., Hur, K., Marks, W.J., Rothlind, J., Sagher, O., Moy, C., Pahwa, R., Burchiel, K., Hogarth, P., Lai, E.C., Duda, J.E., Holloway, K., Samii, A., Horn, S., Bronstein, J.M., Stoner, G., Starr, P.A., Simpson, R., Baltuch, G., Salles, A.D., Huang, G.D., Reda, D.J., 2012. Randomized trial of deep brain stimulation for Parkinson disease: Thirty-six-month outcomes. Neurology 79, 55–65. 10.1212/WNL.0b013e31825dcdc1

